# Describing the population experiencing COVID-19 vaccine breakthrough following second vaccination in England: A cohort study from OpenSAFELY

**DOI:** 10.1101/2021.11.08.21265380

**Authors:** The OpenSAFELY Collaborative:, Amelia Green, Helen Curtis, William Hulme, Elizabeth Williamson, Helen McDonald, Krishnan Bhaskaran, Christopher Rentsch, Anna Schultze, Brian MacKenna, Viyaasan Mahalingasivam, Laurie Tomlinson, Alex Walker, Louis Fisher, Jon Massey, Colm Andrews, Lisa Hopcroft, Caroline Morton, Richard Croker, Jessica Morley, Amir Mehrkar, Seb Bacon, David Evans, Peter Inglesby, George Hickman, Tom Ward, Simon Davy, Rohini Mathur, John Tazare, Rosalind M Eggo, Kevin Wing, Angel Wong, Harriet Forbes, Chris Bates, Jonathan Cockburn, John Parry, Frank Hester, Sam Harper, Ian Douglas, Stephen Evans, Liam Smeeth, Ben Goldacre

## Abstract

**Background:** While the vaccines against COVID-19 are considered to be highly effective, COVID-19 vaccine breakthrough is likely and a small number of people will still fall ill, be hospitalised, or die from COVID-19, despite being fully vaccinated. With the continued increase in numbers of positive SARS-CoV-2 tests, describing the characters of individuals who have experienced a COVID-19 vaccine breakthrough could be hugely important in helping to determine who may be at greatest risk.

**Method:** With the approval of NHS England we conducted a retrospective cohort study using routine clinical data from the OpenSAFELY TPP database of fully vaccinated individuals, linked to secondary care and death registry data, and described the characteristics of those experiencing a COVID-19 vaccine breakthrough.

**Results:** As of 01^st^ November 2021, a total of 15,436,455 individuals were identified as being fully vaccinated against COVID-19, with a median follow-up time of 149 days (IQR: 107-179). From within this population, a total of 577245 (<4%) individuals reported a positive SARS-CoV-2 test. For every 1000 years of patient follow-up time, the corresponding incidence rate was 98.02 (95% CI 97.9-98.15). There were 16,120 COVID-19-related hospital admissions, 1,100 COVID-19 critical care admission patients and 3,925 COVID-19-related deaths; corresponding incidence rates of 2.72 (95% C 2.7-2.74), 0.19 (95% C 0.18-0.19) and 0.66 (95% C 0.65-0.67), respectively. When broken down by the initial priority group, higher rates of hospitalisation and death were seen in those in care homes and those over 80 years of age. Comorbidities with the highest rates of breakthrough COVID-19 included chronic kidney disease, dialysis, transplant, haematological malignancy, and immunocompromised.

**Conclusion:** The majority of COVID-19 vaccine breakthrough cases in England were mild with relatively few fully vaccinated individuals being hospitalised or dying as a result. However, some concerning differences in rates of breakthrough cases were identified in several clinical and demographic groups. While it is important to note that these findings are simply descriptive and cannot be used to answer why certain groups have higher rates of COVID-19 breakthrough than others, the emergence of the Omicron variant of COVID-19 coupled with the continued increase in numbers of positive SARS-CoV-2 tests are concerning. As numbers of fully vaccinated individuals increases and follow-up time lengthens, so too will the number of COVID-19 breakthrough cases. Additional analyses, aimed at identifying individuals at higher risk, are therefore required.

## Background

The vaccination programme for COVID-19 in the United Kingdom (UK) was started on 8 December 2020. Initially, the recipients of COVID-19 vaccines included those in care homes, those over 80 years old and frontline health and social care workers. Those aged ≥70 years or clinically extremely vulnerable were the next priority group, followed by remaining adults in order of decreasing age or at increased risk. As of November 1^st^, 2021 78.7% of individuals aged over 16 in England had been fully vaccinated (i.e., ≥14 days have passed since the receipt of their second dose of a COVID-19 vaccine) [1].

The vaccines against COVID-19 are considered to be highly effective and there is clear evidence that the UK’s COVID-19 vaccination programme has reduced infection and severe outcomes in vaccinated individuals [2, 3]. However, as no vaccine is 100% effective, COVID-19 vaccine breakthrough is likely (i.e., a small number of people will still get sick, be hospitalised, or die from COVID-19, despite being fully vaccinated). Describing individuals who have experienced a COVID-19 vaccine breakthrough could provide the first indication that the COVID-19 vaccine is less effective in certain demographic groups and could be hugely important in helping to determine who may be at greatest risk and therefore might benefit most from booster doses of vaccine.

To that end we used the new secure data analytics platform, OpenSAFELY [4] (built by our group on behalf of NHS England to support analysis of important questions related to COVID-19) to: describe breakthrough COVID-19 among fully vaccinated individuals in England; and to describe how breakthrough COVID-19 varied between priority groups and by clinical and demographic characteristics.

## Methods

### Data sources

This OpenSAFELY study was conducted using the OpenSAFELY-TPP database which contains records for approximately 24 million people currently registered with GP surgeries using TPP SystmOne software (approximately 40% of the English population).

### Study population

The base population consisted of all individuals aged 16 or over with records indicating that they had received two COVID-19 vaccination doses since 8 December 2020 (the earliest vaccination date in England) and who were still alive and registered 2 weeks after their second dose. Individuals were excluded if they tested positive for SARS-CoV-2 or had been hospitalised or died due to COVID-19 within the 2 weeks after their second dose. Follow up started 14 days after an individual’s second dose of the COVID-19 vaccination (the point by which individuals were classified as being fully vaccinated) and individuals were followed up until 01^st^ November 2021 (the latest date for which data were available at the time of the analysis) or until the first occurrence of: the outcome of interest; death; practice de-registration.

### Outcomes

Four outcomes were assessed: positive SARS-CoV-2 swab test, via SGSS and based on swab date (we did not distinguish between symptomatic and asymptomatic infection for this outcome, nor PCR and lateral flow testing); COVID-19 related hospital admissions via HES in-patient records (using both primary and non-primary diagnosis codes); COVID-19 related critical care admissions via HES and COVID-19-related death, via linked death registry records, which included individuals who died within 28-days of positive SARS-CoV-2 test or who had COVID-19 mentioned on the death certificate as one of the causes. COVID-19 hospitalisation variables were derived using ICD-10 COVID-19 diagnosis codes [5]. Outcomes were only included if they occurred 14 days or more after an individual’s second dose of a COVID-19 vaccination.

### Priority groups for vaccination

Individuals were classified into seven priority groups (box 1), based on the Joint Committee on Vaccination and Immunisation (JCVI) priority groups [6] using SNOMED-CT codelists and logic defined in the national COVID-19 Vaccination Uptake Reporting Specification developed by PRIMIS [7]. Individuals were assigned only to their highest priority group and not included again as part of any other priority group. In line with the national reporting specification, most criteria were ascertained using the latest available data at the time of analysis, with the exception of age which was calculated as at 31 March 2021 as recommended by Public Health England.

#### Box 1

***Priority groups for vaccination***.

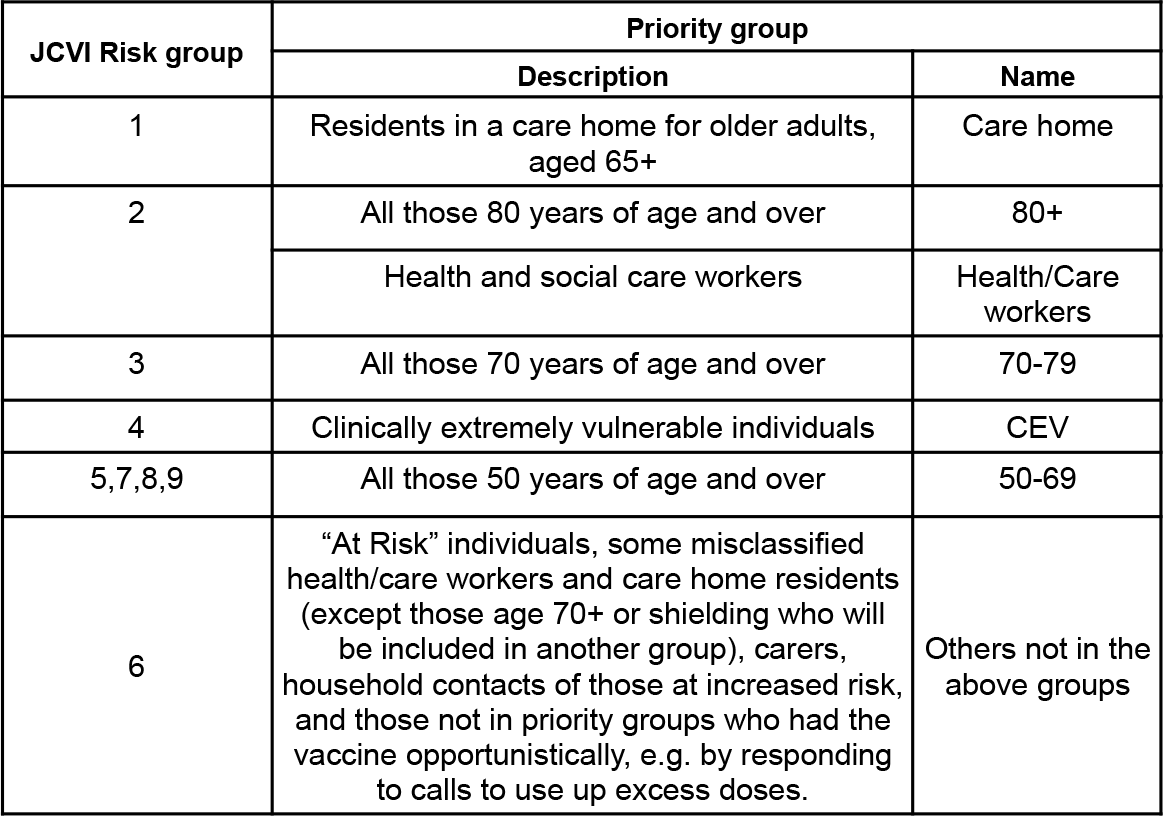

### Key demographic and clinical characteristics

Key clinical and demographic groups which were considered to have a higher possibility of experiencing a COVID-19 vaccine breakthrough were identified from previous studies [8–12]. This included age, sex, body mass index (BMI; kg/m^2^), smoking status, deprivation (measured by the Index of Multiple Deprivation (IMD) as quintiles, ethnicity, NHS region of patient’s general practice, asplenia, asthma, blood pressure, cancer, chronic kidney disease, diabetes mellitus, dialysis, heart disease, haematological malignancy, immunocompromised, learning disability, liver disease, neurological diseases, respiratory disease, severe mental illness and transplant. Other variables considered were time since being fully vaccinated, time between vaccinations and any evidence of a prior SARS-CoV-2 infection.

### Codelists and implementation

Information on all covariates were obtained from primary care records by searching TPP SystmOne records for specific coded data. Detailed information on compilation and sources for every individual codelist is available at https://codelists.opensafely.org/ and the lists are available for inspection and re-use by the broader research community.

### Missing data

Individuals with missing ethnicity, IMD and region were included as “Unknown”.

### Statistical methods

Simple descriptive statistics were used to summarise the counts and rates of COVID-19 vaccine breakthrough. Rates for each outcome were estimated by dividing the count by person-years, with 95% confidence intervals. Counts and rates were stratified by initial priority groups for all outcomes and by selected clinical and demographic groups.

### Software and Reproducibility

Data management and analysis was performed using the OpenSAFELY software libraries, Python 3 and R version 4.0.2. All code for the OpenSAFELY platform for data management, analysis and secure code execution is shared for review and re-use under open licenses at GitHub.com/OpenSAFELY. All code for data management and analysis for this paper is shared for scientific review and re-use under open licenses on GitHub (https://github.com/opensafely/covid-19-vaccine-breakthrough).

### Patient and Public Involvement

Any patient or member of the public is invited to contact us at https://opensafely.org/ regarding this study or the broader OpenSAFELY project.

## Results

### Study population

Out of approximately 24m patients, 15,436,455 individuals were identified as being fully vaccinated against COVID-19 and included in this study (Figure 1). The median follow-up time was 149 days (interquartile range: 107 - 179 days). Since being fully vaccinated, 41% of the base population had at least one record for a (positive/negative) SARS-CoV-2 test recorded, with a positivity rate of 2%. Testing behaviours varied between priority groups with individuals in care homes testing more regularly than other groups; 77% of care home residents had 3+ SARS-CoV-2 tests since being fully vaccinated vs 15%-25% in other groups. The total number of COVID-19 vaccine breakthrough cases was 578,835 (<4%). Table 1 shows the number and rate of individuals fully vaccinated broken down by initial JCVI priority groups, along with the number and rate (per 1,000 patient years) of each outcome.

**Table 1.** Number of individuals fully vaccinated (2 doses + 2 weeks) in initial priority groups in OpenSAFELY-TPP, and number and rate with each outcome.

| Priority Group | Fully vaccinated |  |  |  |  |  | Positive SARS-CoV-2 test |  |  | COVID-19-related hospital admission<sup>†</sup> |  | COVID-19-related critical care admission<sup>†</sup> |  | COVID-19 related death |  |
| --- | --- | --- | --- | --- | --- | --- | --- | --- | --- | --- | --- | --- | --- | --- | --- |
|  | Count* | Follow-up time, medium days (IQR) | Tests conducted (%) |  |  |  | Positivity (%) | Events* (PYs) | Incidence rate per 1000 person-years (95% CI) | Events* (PYs) | Incidence rate per 1000 person-years (95% CI) | Events* (PYs) | Incidence rate per 1000 person-years (95% CI) | Events* (PYs) | Incidence rate per 1000 person-years (95% CI) |
|  |  |  | 0 | 1 | 2 | 3+ |  |  |  |  |  |  |  |  |  |
| All | 15,436,455 | 149 (107-179) | 59 | 17 | 7 | 14 | 2 | 577245 (5888840) | 98.02 (97.9-98.15) | 16120 (5928048) | 2.72 (2.70-2.74) | 1110 (5930338) | 0.19 (0.18-0.19) | 3925 (5972152) | 0.66 (0.65-0.67) |
| Care home (priority group 1) | 98,250 | 200 (180-207) | 12 | 6 | 5 | 77 | 1 | 3105 (48130) | 64.49 (63.35-65.66) | 505 (48301) | 10.46 (10.00-10.93) | [REDACTED] | [REDACTED] | 475 (48595) | 9.75 (9.32-10.21) |
| 80+ (priority group 2) | 1,069,840 | 200 (191-213) | 66 | 13 | 6 | 16 | 2 | 15895 (595480) | 26.69 (26.48-26.9) | 4600 (594247) | 7.74 (7.63-7.85) | 110 (594923) | 0.18 (0.17-0.2) | 1715 (597732) | 2.87 (2.80-2.94) |
| Health/care workers (priority groups 1-2) | 543,005 | 201 (183-211) | 41 | 22 | 11 | 26 | 2 | 29195 (277847) | 105.08 (104.47-105.7) | 235 (281258) | 0.83 (0.78-0.89) | 15 (281298) | 0.06 (0.05-0.08) | 10 (282766) | 0.04 (0.03-0.05) |
| 70-79 (priority groups 3-4) | 1,983,930 | 184 (176-191) | 66 | 15 | 6 | 13 | 2 | 41170 (982158) | 41.92 (41.71-42.13) | 4055 (981792) | 4.13 (4.07-4.20) | 290 (982356) | 0.3 (0.28-0.32) | 980 (987739) | 0.99 (0.96-1.03) |
| CEV (age 16-69) (priority group 4) | 723,215 | 171 (157-181) | 52 | 17 | 8 | 22 | 2 | 34725 (322102) | 107.81 (107.23-108.39) | 2565 (325217) | 7.89 (7.74-8.05) | 335 (325575) | 1.03 (0.97-1.08) | 460 (327565) | 1.40 (1.33-1.47) |
| 50 - 69 (priority groups 5-9) | 4,712,910 | 151 (138-166) | 57 | 17 | 7 | 19 | 2 | 184245 (1939494) | 95 (94.78-95.22) | 2660 (1953510) | 1.36 (1.34-1.39) | 280 (1953880) | 0.14 (0.13-0.15) | 255 (1966741) | 0.13 (0.12-0.14) |
| Others not in the above groups*** | 6,305,305 | 97 (65-136) | 59 | 18 | 8 | 15 | 2 | 268910 (1723630) | 156.01 (155.71-156.32) | 1500 (1743721) | 0.86 (0.84-0.88) | 80 (1743939) | 0.04 (0.04-0.05) | 30 (1761013) | 0.02 (0.01-0.02) |

<sup>†</sup> Calculated as of 30<sup>th</sup> September 2021, due to data availability. Other outcomes calculated as of 01<sup>st</sup> November 2021.
\*All counts (of people and events) have been rounded to the nearest 5 and counts < 8 have been redacted
\*\*This is a count of (positive and negative) test results and may include multiple tests for an individual person
\*\*\*Others includes: others in priority groups ("At Risk"), some misclassified health/care workers and care home residents (except those age 70+ or shielding who will be included in another group), carers, household contacts of those at increased risk, and those not in priority groups who had the vaccine opportunistically, e.g. by responding to calls to use up excess doses.

**Figure 1.**
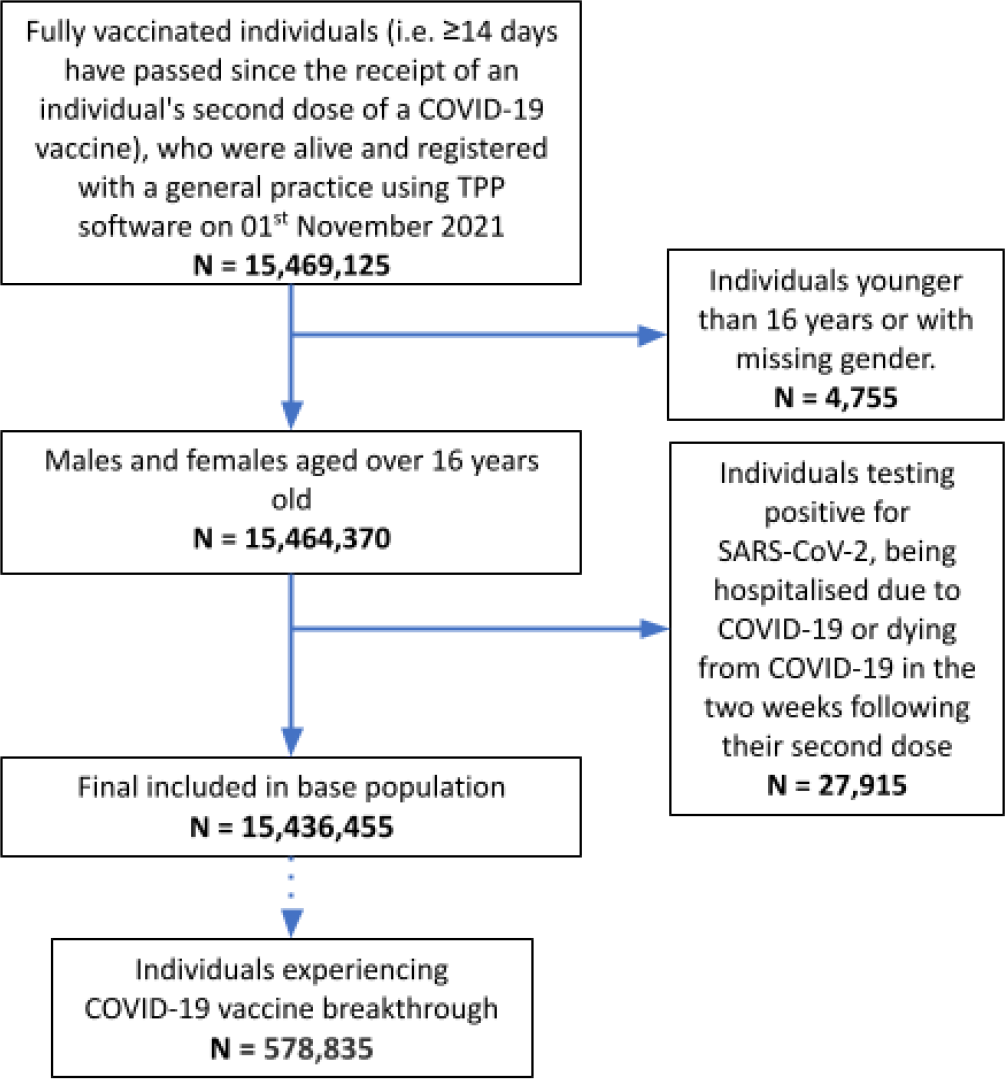
Inclusion/exclusion flowchart

### Positive SARS-CoV-2 test

In fully vaccinated individuals, the median number of days to a positive test for SARS-CoV-2 was 99 (IQR 63 - 134 days) with a total of 577,245 (<4%) individuals testing positive for SARS-CoV-2. For every 1,000 years of patient follow-up time, the corresponding incidence rate was 98.02 (95% CI 97.9-98.15). Within the initial JCVI priority groups, positive SARS-CoV-2 test rates were highest in the CEV group (107.81, 95% CI 107.23-108.39) and lowest in those over 80 years of age (26.69, 95% CI 26.48-26.9). The overall cumulative incidence of positive SARS-CoV-2 tests at 275 days from the study start date was <0.1% (Figure 2).

**Figure 2.**
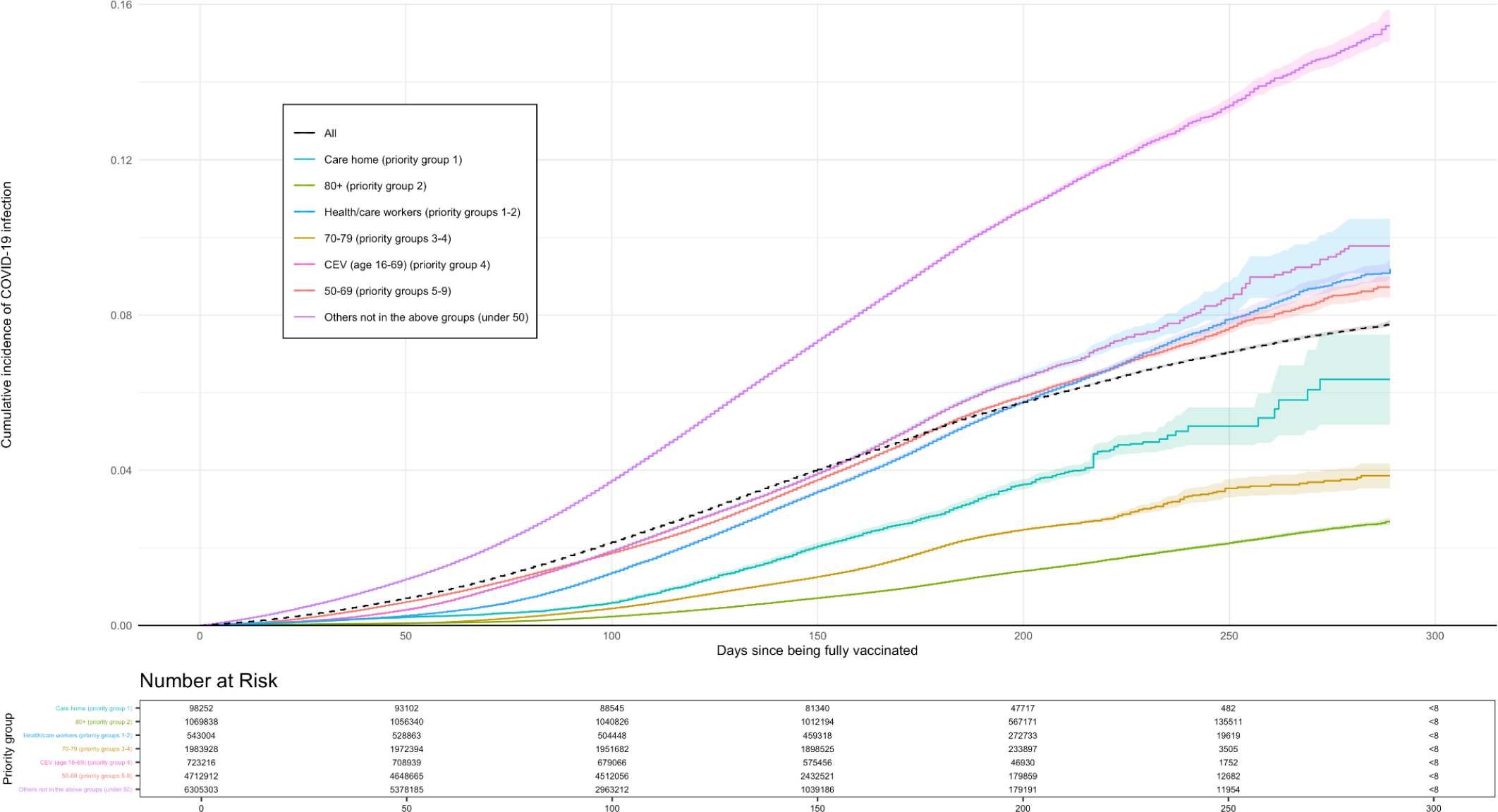
Kaplan-Meir plot for positive SARS-CoV-2 test over time, by priority group

When broken down into clinical/demographic groups and comorbidities (Table 2), rates of individuals testing positive for SARS-CoV-2 were highest in 40-40 year olds (179.93, 95% CI 179.47-180.39) and lowest in individuals over 80 years of age (28.9, 95% CI 28.69-29.12). Rates were higher in females than in males: 102.57 (95% CI 102.39-102.75) vs 92.72 (95% CI 92.54-92.91), respectively. Comorbidities with the highest rates of positive SARS-CoV-2 tests included kidney transplant, dialysis and immunocompromised; 142.92 (95% CI 135.87-150.33), (134.6, 95% CI 124.15-145.93), 91.5 (95% CI 90.88-92.12), respectively. Rates of positive SARS-CoV-2 tests were lowest 0-4 weeks after being fully vaccinated and increased with time since being fully vaccinated: 82.26 (95% CI 79.83-84.77) at 0-4 weeks vs 100.26 (95% CI 100.12-100.39) at 12+ weeks).

**Table 2.** *Count and rates of breakthrough* positive SARS-CoV-2 tests *and hospitalisation and death in fully vaccinated individuals in OpenSAFELY-TPP, broken down by selected clinical and demographic groups*

| Clinical/<br>demographic<br>group | Category | Fully vaccinated |  |  |  |  |  |  | Positive SARS-CoV-2 test |  | Hospitalised with COVID-19 |  | COVID-19-related critical care admission |  | COVID-19 related death |  |
| --- | --- | --- | --- | --- | --- | --- | --- | --- | --- | --- | --- | --- | --- | --- | --- | --- |
|  |  | Count* | Follow-up time, medium days (IQR) | Tests conducted (%) |  |  |  | Positivity (%) | Events* (PYs) | Incidence rate per 1000 person-years (95% CI) | Events* (PYs) | Incidence rate per 1000 person-years (95% CI) | Events* (PYs) | Incidence rate per 1000 person-years (95% CI) | Events* (PYs) | Incidence rate per 1000 person-years (95% CI) |
|  |  |  |  | 0 | 1 | 2 | 3+ |  |  |  |  |  |  |  |  |  |
| Age | 16 - 29 | 2214530 | 66 (47-129) | 63 | 17 | 7 | 13 | 2.29 | 63305 (513316) | 123.33 (122.84-123.82) | 420 (518128) | 0.81 (0.77-0.85) | 20 (518198) | 0.04 (0.03-0.05) | [REDACTED] | [REDACTED] |
|  | 30 - 39 | 2277680 | 94 (72-145) | 58 | 19 | 8 | 15 | 2.42 | 92860 (657013) | 141.34 (140.87-141.8) | 705 (663471) | 1.06 (1.02-1.1) | 45 (663571) | 0.06 (0.06-0.08) | 20 (669753) | 0.03 (0.02-0.04) |
|  | 40 - 50 | 2448735 | 126 (103-155) | 53 | 19 | 8 | 20 | 2.61 | 152530 (847728) | 179.93 (179.47-180.39) | 1185 (861243) | 1.37 (1.34-1.42) | 115 (861409) | 0.13 (0.12-0.15) | 65 (868091) | 0.07 (0.06-0.08) |
|  | 50 - 59 | 2930705 | 146 (135-163) | 54 | 17 | 8 | 21 | 1.74 | 135950 (1184298) | 114.8 (114.48-115.11) | 2040 (1196666) | 1.71 (1.67-1.74) | 230 (1196963) | 0.19 (0.18-0.21) | 190 (1204969) | 0.16 (0.15-0.17) |
|  | 60 - 69 | 2412600 | 163 (150-174) | 60 | 16 | 7 | 18 | 1.47 | 72445 (1060342) | 68.32 (68.07-68.57) | 2635 (1063843) | 2.48 (2.43-2.53) | 295 (1064197) | 0.28 (0.26-0.3) | 490 (1070781) | 0.46 (0.44-0.48) |
|  | 70 - 79 | 2009505 | 184 (176-191) | 65 | 15 | 6 | 14 | 1.84 | 41915 (995030) | 42.13 (41.92-42.33) | 4170 (994697) | 4.19 (4.13-4.26) | 295 (995277) | 0.3 (0.28-0.31) | 1035 (1000725) | 1.03 (1-1.07) |
|  | 80 + | 1142695 | 200 (191-213) | 63 | 12 | 6 | 20 | 1.41 | 18240 (631113) | 28.9 (28.69-29.12) | 4970 (630019) | 7.89 (7.77-8) | 115 (630741) | 0.18 (0.16-0.2) | 2125 (633714) | 3.35 (3.28-3.43) |
| Sex | Female | 8038785 | 155 (117-184) | 55 | 17 | 8 | 20 | 1.69 | 325170 (3170186) | 102.57 (102.39-102.75) | 7455 (3194555) | 2.33 (2.31-2.36) | 410 (3195650) | 0.13 (0.12-0.13) | 1600 (3217391) | 0.50 (0.49-0.51) |
|  | Male | 7397670 | 144 (99-174) | 63 | 17 | 7 | 14 | 2.57 | 252080 (2718654) | 92.72 (92.54-92.91) | 8670 (2733512) | 3.17 (3.14-3.21) | 705 (2734707) | 0.26 (0.25-0.27) | 2325 (2754761) | 0.84 (0.83-0.86) |
| BMI (kg/m2) | Not obese | 12072875 | 145 (98-178) | 59 | 17 | 7 | 17 | 1.94 | 429525 (4496521) | 95.52 (95.38-95.67) | 10230 (4524028) | 2.26 (2.24-2.28) | 580 (4525512) | 0.13 (0.12-0.13) | 2735 (4558129) | 0.60 (0.59-0.61) |
|  | Obese (>30) | 3363580 | 159 (135-181) | 57 | 17 | 7 | 18 | 2.14 | 147725 (1392319) | 106.1 (105.82-106.37) | 5895 (1404038) | 4.2 (4.14-4.25) | 535 (1404845) | 0.38 (0.36-0.4) | 1190 (1414023) | 0.84 (0.82-0.87) |
| Smoking | Non-smoker and unknown | 7798320 | 143 (91-176) | 58 | 17 | 7 | 18 | 1.93 | 289690 (2823672) | 102.59 (102.4-102.78) | 5470 (2843372) | 1.92 (1.9-1.95) | 360 (2844181) | 0.13 (0.12-0.13) | 1110 (2865276) | 0.39 (0.38-0.4) |
|  | Current and former | 7638130 | 156<br>(125-182) | 59 | 17 | 7 | 17 | 2.04 | 287555<br>(3065168) | 93.81<br>(93.64-93.99) | 10650<br>(3084695) | 3.45<br>(3.42-3.49) | 750<br>(3086176) | 0.24<br>(0.23-0.25) | 2815<br>(3106876) | 0.91<br>(0.89-0.92) |
| Ethnicity | White | 11775235 | 154<br>(119-182) | 57 | 17 | 8 | 18 | 1.98 | 461300<br>(4644112) | 99.33<br>(99.18-99.48) | 14385<br>(4676873) | 3.08<br>(3.05-3.1) | 970<br>(4678913) | 0.21<br>(0.2-0.21) | 3595<br>(4710788) | 0.76<br>(0.75-0.78) |
|  | Mixed | 149745 | 130<br>(77-164) | 59 | 17 | 8 | 16 | 1.94 | 4950 (49600) | 99.82<br>(98.41-101.25) | 100<br>(49930) | 1.96<br>(1.77-2.17) | 10<br>(49943) | 0.18<br>(0.13-0.25) | 10<br>(50348) | 0.24<br>(0.18-0.32) |
|  | Asian or Asian British | 893395 | 135<br>(82-170) | 67 | 16 | 6 | 11 | 2.78 | 29590<br>(306860) | 96.43<br>(95.87-96.99) | 1050<br>(308624) | 3.4<br>(3.29-3.5) | 85<br>(308779) | 0.27<br>(0.24-0.3) | 200<br>(311202) | 0.64<br>(0.6-0.69) |
|  | Black or Black British | 222800 | 138<br>(87-172) | 62 | 15 | 7 | 16 | 1.39 | 5870 (78070) | 75.2<br>(74.23-76.19) | 215<br>(78347) | 2.76<br>(2.58-2.95) | 15<br>(78378) | 0.2<br>(0.16-0.26) | 30<br>(78981) | 0.38<br>(0.32-0.46) |
|  | Other ethnic groups | 239330 | 123<br>(79-159) | 63 | 17 | 7 | 13 | 1.94 | 5885 (77821) | 75.6<br>(74.62-76.59) | 150<br>(77983) | 1.94<br>(1.78-2.1) | 15<br>(78005) | 0.19<br>(0.15-0.25) | 30<br>(78651) | 0.39<br>(0.33-0.47) |
|  | Unknown | 2155945 | 135<br>(83-164) | 66 | 15 | 6 | 14 | 1.90 | 69655<br>(732379) | 95.11<br>(94.75-95.47) | 220<br>(736309) | 0.3<br>(0.28-0.32) | 15<br>(736339) | 0.02<br>(0.02-0.03) | 60<br>(742182) | 0.08<br>(0.07-0.09) |
| IMD quintile | 1 (most deprived) | 2511445 | 146<br>(100-176) | 65 | 15 | 6 | 14 | 2.33 | 96345<br>(924825) | 104.18<br>(103.84-104.51) | 4070<br>(932356) | 4.37<br>(4.3-4.44) | 270<br>(932945) | 0.29<br>(0.27-0.31) | 965<br>(939760) | 1.02<br>(0.99-1.06) |
|  | 2 | 2859260 | 148<br>(101-178) | 61 | 16 | 7 | 16 | 2.02 | 104825<br>(1070325) | 97.94<br>(97.64-98.24) | 3260<br>(1077574) | 3.03<br>(2.97-3.08) | 235<br>(1078034) | 0.22<br>(0.21-0.23) | 775<br>(1085778) | 0.71<br>(0.69-0.74) |
|  | 3 | 3304590 | 150<br>(109-180) | 59 | 17 | 7 | 17 | 1.89 | 119140<br>(1268886) | 93.89<br>(93.62-94.17) | 3145<br>(1276431) | 2.46<br>(2.42-2.51) | 220<br>(1276870) | 0.17<br>(0.16-0.18) | 780<br>(1285811) | 0.61<br>(0.59-0.63) |
|  | 4 | 3259125 | 151<br>(113-181) | 57 | 17 | 8 | 18 | 1.92 | 121195<br>(1267010) | 95.65<br>(95.38-95.93) | 2900<br>(1274776) | 2.28<br>(2.23-2.32) | 190<br>(1275184) | 0.15<br>(0.14-0.16) | 725<br>(1284002) | 0.56<br>(0.54-0.59) |
|  | 5 (least deprived) | 3101015 | 152<br>(116-181) | 54 | 18 | 8 | 19 | 1.88 | 119905<br>(1215555) | 98.64<br>(98.36-98.93) | 2365<br>(1223512) | 1.93<br>(1.89-1.97) | 165<br>(1223851) | 0.14<br>(0.13-0.15) | 600<br>(1232240) | 0.49<br>(0.47-0.51) |
|  | Unknown | 401020 | 139<br>(87-173) | 57 | 18 | 8 | 17 | 2.19 | 15840<br>(142239) | 111.35<br>(110.47-112.24) | 380 (143418) | 2.66<br>(2.53-2.8) | 25<br>(143473) | 0.19<br>(0.16-0.23) | 80<br>(144561) | 0.55<br>(0.49-0.62) |
| Region | London | 890235 | 129<br>(84-170) | 55 | 19 | 9 | 17 | 1.81 | 21930<br>(304549) | 72.01<br>(71.52-72.5) | 585<br>(305425) | 1.92<br>(1.84-2) | 40<br>(305507) | 0.13<br>(0.11-0.15) | 135<br>(307899) | 0.44<br>(0.40-0.47) |
|  | East of England | 3614790 | 149<br>(104-179) | 57 | 17 | 7 | 18 | 1.56 | 113155<br>(1374549) | 82.32<br>(82.08-82.57) | 2610<br>(1379657) | 1.89<br>(1.85-1.93) | 200<br>(1380037) | 0.15<br>(0.14-0.16) | 700<br>(1389826) | 0.51<br>(0.49-0.52) |
|  | East Midlands | 2695775 | 150<br>(113-177) | 60 | 16 | 7 | 16 | 2.19 | 107955<br>(1027450) | 105.07<br>(104.75-105.39) | 3130<br>(1035106) | 3.02<br>(2.97-3.08) | 255<br>(1035520) | 0.25<br>(0.23-0.26) | 715<br>(1042843) | 0.69<br>(0.66-0.71) |
|  | North East | 744765 | 150<br>(115-181) | 60 | 17 | 7 | 15 | 3.23 | 37560<br>(289183) | 129.88<br>(129.21-130.55) | 1285<br>(292567) | 4.4<br>(4.27-4.52) | 80<br>(292754) | 0.27<br>(0.24-0.31) | 300<br>(294778) | 1.01<br>(0.96-1.07) |
|  | North West | 1388335 | 150<br>(115-180) | 60 | 16 | 7 | 16 | 2.48 | 63060<br>(536308) | 117.59<br>(117.12-118.05) | 2035<br>(542248) | 3.75<br>(3.67-3.84) | 155<br>(542552) | 0.29<br>(0.27-0.31) | 500 (546324) | 0.92<br>(0.88-0.96) |
|  | South East | 1076540 | 151<br>(109-180) | 57 | 17 | 7 | 18 | 1.56 | 34605<br>(416926) | 83.01<br>(82.56-83.45) | 850<br>(418738) | 2.03<br>(1.96-2.1) | 55<br>(418859) | 0.13<br>(0.11-0.15) | 210<br>(421760) | 0.50<br>(0.46-0.53) |
|  | South West | 2311580 | 152<br>(111-180) | 57 | 17 | 7 | 19 | 1.69 | 82950<br>(895885) | 92.59<br>(92.27-92.91) | 1845<br>(900478) | 2.05<br>(2-2.1) | 100<br>(900722) | 0.11<br>(0.1-0.12) | 445<br>(906965) | 0.49<br>(0.47-0.51) |
|  | West Midlands | 540825 | 149<br>(109-179) | 63 | 15 | 6 | 15 | 2.44 | 21005<br>(205943) | 102<br>(101.3-102.71) | 860<br>(207667) | 4.13<br>(3.99-4.28) | 55<br>(207805) | 0.26<br>(0.23-0.3) | 190<br>(209262) | 0.91<br>(0.84-0.98) |
|  | Yorkshire and<br>The Humber | 2164945 | 151<br>(111-181) | 61 | 16 | 7 | 16 | 2.40 | 94755<br>(835263) | 113.44<br>(113.08-113.81) | 2915<br>(843381) | 3.46<br>(3.39-3.52) | 170<br>(843799) | 0.20<br>(0.19-0.22) | 730<br>(849672) | 0.86<br>(0.83-0.89) |
|  | Unknown | 8665 | 122<br>(70-166) | 59 | 18 | 7 | 16 | 1.99 | 270 (2784) | 96.97<br>(91.25-103.06) | [REDACTED] | [REDACTED] | [REDACTED] | [REDACTED] | [REDACTED] | [REDACTED] |
| Asplenia |  | 107930 | 170<br>(154-186) | 51 | 18 | 9 | 22 | 1.88 | 4865 (48811) | 99.65<br>(98.23-101.09) | 160<br>(49242) | 3.29<br>(3.04-3.56) | 10<br>(49267) | 0.2<br>(0.15-0.28) | 35<br>(49558) | 0.71<br>(0.60-0.84) |
| Asthma |  | 2325235 | 151<br>(110-178) | 55 | 18 | 8 | 19 | 2.21 | 104500<br>(889055) | 117.54<br>(117.18-117.91) | 3355<br>(897673) | 3.74<br>(3.68-3.8) | 235<br>(898153) | 0.26<br>(0.25-0.28) | 650<br>(904463) | 0.72<br>(0.69-0.75) |
| Blood pressure | Normal | 3176645 | 137<br>(87-171) | 55 | 18 | 8 | 19 | 2.04 | 135525<br>(1108305) | 122.28<br>(121.95-122.62) | 2745<br>(1118981) | 2.45<br>(2.41-2.5) | 160<br>(1119372) | 0.14<br>(0.13-0.15) | 745<br>(1127941) | 0.66<br>(0.63-0.68) |
|  | Elevated/high | 3415235 | 151<br>(122-179) | 58 | 17 | 7 | 18 | 2.00 | 135635<br>(1342391) | 101.04<br>(100.77-101.31) | 3675<br>(1352132) | 2.72<br>(2.67-2.76) | 275<br>(1352657) | 0.2<br>(0.19-0.21) | 875<br>(1361920) | 0.64<br>(0.62-0.66) |
|  | Unknown | 8844575 | 152<br>(113-181) | 60 | 16 | 7 | 16 | 1.96 | 306090<br>(3438143) | 89.03<br>(88.87-89.19) | 9700<br>(3456954) | 2.81<br>(2.78-2.83) | 680<br>(3458328) | 0.2 (0.19-0.2) | 2310<br>(3482291) | 0.66<br>(0.65-0.68) |
| Cancer (non-haematological) |  | 888120 | 180<br>(160-194) | 54 | 16 | 8 | 21 | 1.60 | 25510<br>(424205) | 60.13<br>(59.76-60.51) | 2435<br>(425148) | 5.73<br>(5.62-5.85) | 135<br>(425490) | 0.32<br>(0.29-0.35) | 815<br>(427852) | 1.91<br>(1.84-1.98) |
| Chronic kidney<br>disease | Stage 3a | 527255 | 187<br>(173-201) | 61 | 14 | 6 | 18 | 1.80 | 12135<br>(267915) | 45.29<br>(44.88-45.7) | 2040<br>(267918) | 7.61<br>(7.44-7.78) | 105<br>(268206) | 0.4<br>(0.36-0.44) | 690<br>(269616) | 2.55<br>(2.46-2.65) |
|  | Stage 3b | 205755 | 192<br>(178-205) | 60 | 13 | 6 | 21 | 1.77 | 4440<br>(106744) | 41.59<br>(40.97-42.21) | 1270<br>(106641) | 11.89<br>(11.56-12.23) | 55<br>(106814) | 0.51<br>(0.45-0.59) | 500<br>(107352) | 4.67<br>(4.46-4.88) |
|  | Stage 4 | 51035 | 191<br>(174-204) | 55 | 12 | 7 | 26 | 1.77 | 1275<br>(25816) | 49.35<br>(47.99-50.75) | 465<br>(25788) | 18.11<br>(17.29-18.97) | 25<br>(25846) | 1.04<br>(0.86-1.27) | 215<br>(25976) | 8.28<br>(7.73-8.86) |
|  | Stage 5 | 5505 | 184<br>(164-200) | 37 | 12 | 8 | 44 | 1.42 | 215<br>(2607) | 82.86<br>(77.41-88.69) | 80<br>(2615) | 30.97<br>(27.72-34.61) | 10<br>(2625) | 3.43<br>(2.46-4.78) | 30<br>(2639) | 11.37<br>(9.47-13.65) |
| Diabetes |  | 1471735 | 173<br>(157-191) | 60 | 15 | 7 | 18 | 2.18 | 52365<br>(681398) | 76.85<br>(76.52-77.19) | 5115<br>(684421) | 7.48<br>(7.37-7.58) | 415<br>(685126) | 0.6<br>(0.58-0.63) | 1385<br>(689117) | 2.01<br>(1.96-2.07) |
| Dialysis | Previous kidney transplant | 2335 | 182<br>(172-198) | 33 | 15 | 8 | 43 | 1.75 | 155 (1137) | 134.6<br>(124.15-145.93) | 45<br>(1148) | 39.2<br>(33.77-45.51) | 15<br>(1153) | 12.14<br>(9.29-15.86) | 15<br>(1161) | 14.65<br>(11.49-18.67) |
|  | No previous kidney transplant | 10460 | 180<br>(165-199) | 28 | 9 | 6 | 56 | 1.03 | 530<br>(4957) | 107.13<br>(102.58-111.88) | 130<br>(4997) | 26.22<br>(24.02-28.61) | 20<br>(5016) | 3.59<br>(2.84-4.54) | 40<br>(5044) | 8.33<br>(7.14-9.72) |
| Heart disease |  | 1739390 | 180<br>(163-194) | 58 | 15 | 7 | 20 | 1.86 | 51555<br>(841304) | 61.28<br>(61.01-61.55) | 6370<br>(843257) | 7.56<br>(7.46-7.65) | 375<br>(844154) | 0.45<br>(0.42-0.47) | 2120<br>(848806) | 2.50<br>(2.45-2.55) |
| Haematological malignancy |  | 103190 | 180<br>(166-193) | 52 | 16 | 8 | 24 | 2.27 | 3700<br>(49726) | 74.45<br>(73.23-75.68) | 680<br>(49900) | 13.61<br>(13.09-14.14) | 90<br>(49988) | 1.76<br>(1.58-1.96) | 255<br>(50270) | 5.11<br>(4.8-5.44) |
| Immunocompromised |  | 507050 | 177<br>(159-191) | 52 | 17 | 8 | 22 | 2.21 | 21605<br>(236129) | 91.5<br>(90.88-92.12) | 2225<br>(237772) | 9.36<br>(9.17-9.56) | 275<br>(238055) | 1.16<br>(1.1-1.24) | 745<br>(239440) | 3.10<br>(2.99-3.22) |
| Learning disability |  | 102540 | 160<br>(146-177) | 55 | 11 | 6 | 28 | 1.26 | 3225<br>(43815) | 73.58<br>(72.3-74.89) | 170<br>(43991) | 3.86<br>(3.58-4.17) | 15<br>(44016) | 0.34<br>(0.26-0.44) | 25<br>(44295) | 0.61<br>(0.5-0.74) |
| Liver disease |  | 347125 | 164<br>(144-182) | 56 | 17 | 8 | 19 | 2.22 | 13950<br>(150851) | 92.47<br>(91.69-93.26) | 950<br>(151858) | 6.27<br>(6.07-6.48) | 75<br>(151988) | 0.51<br>(0.45-0.57) | 220<br>(152931) | 1.44<br>(1.34-1.54) |
| Neurological disease |  | 870985 | 176<br>(156-193) | 55 | 15 | 7 | 24 | 1.71 | 28300<br>(406143) | 69.68<br>(69.26-70.09) | 2955<br>(407619) | 7.25<br>(7.11-7.38) | 140<br>(408043) | 0.35<br>(0.32-0.38) | 1040<br>(410355) | 2.53<br>(2.46-2.61) |
| Respiratory disease |  | 579305 | 180<br>(166-193) | 59 | 15 | 7 | 19 | 2.05 | 16700<br>(281045) | 59.41<br>(58.96-59.88) | 2965<br>(281552) | 10.53<br>(10.34-10.72) | 180<br>(281949) | 0.63<br>(0.59-0.68) | 945<br>(283493) | 3.34<br>(3.23-3.45) |
| Severe mental illness |  | 181205 | 159<br>(141-177) | 60 | 14 | 6 | 20 | 1.71 | 5105<br>(75928) | 67.23<br>(66.3-68.18) | 350<br>(76168) | 4.62<br>(4.38-4.87) | 30<br>(76216) | 0.38<br>(0.32-0.46) | 90<br>(76707) | 1.17<br>(1.06-1.3) |
| Transplant | Kidney (with previous dialysis) | 5725 | 179<br>(172-191) | 46 | 17 | 9 | 28 | 3.67 | 390<br>(2736) | 142.92<br>(135.87-150.33) | 115<br>(2762) | 40.91<br>(37.23-44.94) | 30<br>(2775) | 11.53<br>(9.66-13.76) | 30<br>(2794) | 11.45<br>(9.6-13.67) |
|  | Kidney (without previous dialysis) | 4510 | 179<br>(171-191) | 47 | 19 | 9 | 26 | 3.60 | 275<br>(2157) | 128.4<br>(120.91-136.35) | 70<br>(2177) | 33.07<br>(29.4-37.21) | 20<br>(2186) | 8.23<br>(6.5-10.42) | 20<br>(2200) | 10<br>(8.08-12.38) |
|  | Other organ | 6760 | 178<br>(170-188) | 45 | 17 | 10 | 28 | 3.12 | 405<br>(3192) | 126.86<br>(120.71-133.33) | 100<br>(3220) | 31.67<br>(28.69-34.97) | 25<br>(3232) | 7.42<br>(6.05-9.11) | 35<br>(3252) | 10.45<br>(8.81-12.41) |
| Time since being fully vaccinated | 0-4 weeks | 329735 | 16 (8-23) | 86 | 8 | 2 | 3 | 1.24 | 1110<br>(13494) | 82.26<br>(79.83-84.77) | 65<br>(12740) | 4.95<br>(4.36-5.61) | [REDACTED] | [REDACTED] | 70<br>(13507) | 5.26<br>(4.67-5.92) |
|  | 4 - 8 weeks | 882060 | 44 (37-51) | 79 | 12 | 3 | 5 | 1.40 | 5115<br>(105031) | 48.68<br>(48.01-49.37) | 160<br>(102947) | 1.53<br>(1.42-1.66) | 15<br>(102953) | 0.14<br>(0.1-0.18) | 115<br>(105257) | 1.09<br>(1.00-1.2) |
|  | 8 - 12 weeks | 1423100 | 69 (62-76) | 68 | 18 | 6 | 8 | 1.91 | 19485<br>(268969) | 72.45<br>(71.93-72.97) | 335<br>(266644) | 1.25<br>(1.19-1.32) | 50<br>(266662) | 0.19<br>(0.16-0.22) | 255<br>(270469) | 0.94<br>(0.89-1.00) |
|  | 12+ weeks | 12801565 | 159<br>(135-184) | 56 | 17 | 8 | 19 | 2.00 | 551540<br>(5501347) | 100.26<br>(100.12-100.39) | 15565<br>(5545736) | 2.81<br>(2.78-2.83) | 1045<br>(5548002) | 0.19<br>(0.18-0.19) | 3485<br>(5582919) | 0.62<br>(0.61-0.63) |
| Time between vaccinations | 6 weeks or less | 493295 | 187<br>(112-283) | 50 | 19 | 9 | 22 | 1.39 | 17600<br>(251616) | 69.95<br>(69.43-70.48) | 1140<br>(253095) | 4.51<br>(4.38-4.65) | 60<br>(253311) | 0.23<br>(0.2-0.27) | 315<br>(254632) | 1.24<br>(1.17-1.31) |
|  | 6-12 weeks | 13514270 | 150<br>(108-179) | 58 | 17 | 7 | 17 | 2.02 | 517360<br>(5142350) | 100.61<br>(100.47-100.75) | 13040<br>(5178048) | 2.52<br>(2.5-2.54) | 920<br>(5179860) | 0.18<br>(0.17-0.18) | 3065<br>(5216478) | 0.59<br>(0.58-0.6) |
|  | 12 weeks or more | 1428885 | 143<br>(100-170) | 65 | 14 | 6 | 15 | 1.96 | 42290<br>(494874) | 85.45<br>(85.04-85.87) | 1940<br>(496924) | 3.90<br>(3.81-3.99) | 135<br>(497185) | 0.27<br>(0.25-0.3) | 545<br>(501041) | 1.09<br>(1.04-1.13) |
| Prior COVID-19 | Between 1st and 2nd vaccination | 190400 | 65 (42-166) | 64 | 14 | 5 | 17 | 0.38 | 1705<br>(49275) | 34.62<br>(33.79-35.47) | 110<br>(49060) | 2.28<br>(2.08-2.51) | [REDACTED] | [REDACTED] | 45<br>(49596) | 0.93<br>(0.8-1.07) |
|  | Before 1st vaccination | 922025 | 138<br>(88-170) | 55 | 17 | 7 | 21 | 0.35 | 8815<br>(325345) | 27.1<br>(26.81-27.39) | 420<br>(324287) | 1.3<br>(1.24-1.36) | 20<br>(324363) | 0.06<br>(0.05-0.08) | 150 (326845) | 0.45<br>(0.42-0.49) |

### COVID-19 related hospital admission

From within the fully vaccinated population, 16,120 had a COVID-19 related hospital admission (about 1 in 1,000 or 1%), with a median time to hospitalisation of 116 days (IQR 83-148). Of those who had a COVID-19 related hospital admission, 14,740 (91%) had a positive SARS-CoV-2 test in their records with 7,015 (48%) occurring prior to an individual being hospitalised, 5770 (75%) occurring within the first 2 days of their hospital admission, 1795 (23%) between 3 and 29 days after being hospitalised and 160 (2%) occurring 30 days or more after an individual had been hospitalised.

Rates of COVID-19 related hospital admissions increased with age; 0.81 (0.77-0.85) for 16-29 year olds vs 7.89 (7.77-8.00) for those over 80 years, respectively. Rates were higher in those who were more deprived; most deprived IMD quintile versus least deprived IMD quintile was 4.37 (4.3-4.44) vs 1.93 (1.89-1.97), respectively. Comorbidities with the highest rates of COVID-19 related hospital admissions included chronic kidney disease, dialysis and kidney transplant; 30.97 (27.72-34.61), 39.2 (33.77-45.51), 40.91 (37.23-44.94), respectively.

### COVID-19 related critical care admission

Of the 16,120 COVID-19 related hospital admission, 1,110 needed to be admitted to critical care (about 1 in 14 or 7%), with a median time to critical care admission of 104 days (IQR 73-134). For every 1,000 years of patient follow-up time, the corresponding incidence rate was 0.19 (95% CI 0.18-0.19). Within the initial JCVI priority groups, COVID-19 related critical care admissions rates were highest in the CEV group (1.03, 95% CI 0.97-1.08) and lowest in health care workers 0.06 (95% CI 0.05-0.08).

Rates of COVID-19 related critical care admissions were higher in males than females and in; 0.26 (95% CI 0.25-0.27) vs 0.13 (95% CI 0.12-0.13), respectively. Rates were higher in those who were obese compared to not obese; BMI >30 (kg/m2) versus BMI <30 (kg/m2) was 0.38 (0.36-0.4) vs 0.13 (0.12-0.13), respectively. Comorbidities with the highest rates of COVID-19 related critical care admissions included dialysis, kidney transplant and chronic kidney disease; 12.14 (95% CI 9.29-15.86), 11.53 (95% CI 9.66-13.76) and 3.43 (95% CI 2.46-4.78), respectively.

### COVID-19 related death

Of those who were fully vaccinated, 3,925 had a COVID-19 related death (about 1 in 4000 or <0.03%) with a median number of days to death of 143 (IQR 112-173). While the majority (44%) of deaths occurred in the 80+ JCVI priority group, COVID-19 related death rates were over three times as high among individuals living in care homes compared to those over 80 years living in private residences; 9.75 (95% CI 9.32-10.21) vs 2.87 (95% CI 2.8-2.94), respectively.

There were very few deaths in those under 30 years of age with rates of COVID-19-related death increasing with increased age; 0.03 (95% CI 0.02-0.04) for 30-39 year olds vs 3.35 (95% CI 3.28-3.43) for those over 80 years, respectively. Comorbidities with the highest rates of COVID-19-related death included dialysis, transplant and chronic kidney disease; 14.65 (95% CI 11.49-18.67), 11.45 (95% CI 9.60-13.67) and 11.37 (95% CI 9.47-13.65), respectively.

## Discussion

### Summary

This descriptive analysis in over 10 million people living in England shows that COVID-19 vaccine breakthrough is occurring, however, events are currently rare and mostly mild in nature. Nevertheless, some individuals are experiencing higher rates of serious illness and death due to COVID-19, such as those in care homes, with chronic kidney disease, on dialysis, with a transplant or who are immunosuppressed. It is important to note that this analysis is a simple descriptive piece of analysis. While it is possible to adjust rates (as demonstrated for patients with chronic kidney disease in Supplementary Table S1) to help inform decisions around rollout of vaccine/booster programme for patients at high risk of adverse outcomes, it is important to stress that this study was not designed to estimate risk factors for COVID-19 vaccine breakthrough and thus cannot be used to answer why certain groups have higher rates of breakthrough cases than others or to estimate vaccine effectiveness.

### Strengths and weaknesses

This study used large-scale, routinely-collected primary care records, linked to coronavirus testing surveillance, hospital, and death registry data. This allowed us to describe a substantial proportion of the English population in rich longitudinal detail and to detect variations in COVID-19 vaccine breakthrough cases, as early as possible.

We acknowledge several important limitations of these findings. First, even though the base population consisted of over 10 million fully vaccinated individuals the numbers of COVID-19 vaccine breakthrough cases were relatively small, especially for hospitalisations and deaths. This made comparisons between outcomes, specifically at selected clinical and demographic levels difficult,meaning rates could be imprecisely estimated. Second, due to the targeted roll out of the COVID-19 vaccination programme in England, this cohort represents mostly older and vulnerable populations. In addition, follow-up time is systematically different amongst individuals included in this study and no adjustment for this has been made. Third, asymptomatic testing patterns vary between individuals. Apart from health and care workers, asymptomatic individuals without any underlying health issues or comorbidities are less likely to get tested than those with underlying health issues or comorbidities (i.e., haemodialysis patients) who will undergo asymptomatic testing regularly. Most lateral flow tests conducted at home are not included in our data. The number of reported positive SARS-CoV-2 tests is therefore likely to be an undercount of SARS-CoV-2 among fully vaccinated individuals without any underlying health issues or comorbidities, which may have led to underestimation of the corresponding rates. Fourth, characteristics linked to COVID-19 vaccine breakthrough in fully vaccinated individuals may be reflective of higher infection rates regardless of vaccination in some groups, not because of vaccination (i.e. due to higher exposure due to behavioural differences) [13].

### Findings in Context

Although COVID-19 vaccinations have demonstrated safety and efficacy in healthy volunteers [14], some people still contract COVID-19 after vaccination and are at risk of serious COVID-19 outcomes (in particular, hospital admission or death). However, with only a handful of studies investigating risk factors for post-vaccination infection [8, 11, 12], very little is known about how breakthrough COVID-19 varies between key clinical and demographic groups. Our findings on COVID-19 vaccine breakthrough are consistent with patterns observed worldwide; COVID-19 vaccine breakthrough cases are rare and mild. However, there are potentially several groups who are at higher risk of COVID-19 vaccine breakthrough including those in care-homes, with chronic kidney disease, on dialysis, with transplants or who are immunocompromised.

### Conclusion

As of 1^st^ November 2021, the majority of COVID-19 vaccine breakthrough cases in England were mild with relatively smaller numbers of fully vaccinated individuals being hospitalised or dying as a result of COVID-19. While these numbers are in line with expectations, and while follow-up time for fully vaccinated individuals is limited and variation in vaccination coverage between groups and regions will have many complex drivers, some differences in rates of breakthrough cases were identified in several clinical and demographic groups. The emergence of the Omicron variant of COVID-19 coupled with the continued increase in numbers of positive SARS-CoV-2 tests and as numbers of fully vaccinated individuals increases and follow-up time lengthens, so too will the number of COVID-19 breakthrough cases. Additional analyses are therefore needed to enable identification of individuals at higher risk, who would require continued strict precautions, and additional vaccination.

## Data Availability

Access to the underlying identifiable and potentially re-identifiable pseudonymised electronic health record data is tightly governed by various legislative and regulatory frameworks, and restricted by best practice. The data in OpenSAFELY is drawn from General Practice data across England where TPP is the Data Processor. TPP developers (CB, RC, JP, FH, and SH) initiate an automated process to create pseudonymised records in the core OpenSAFELY database, which are copies of key structured data tables in the identifiable records. These are linked onto key external data resources that have also been pseudonymised via SHA-512 one-way hashing of NHS numbers using a shared salt. DataLab developers and PIs (BG, LS, CEM, SB, AJW, WH, DE, PI, and CTR) holding contracts with NHS England have access to the OpenSAFELY pseudonymised data tables as needed to develop the OpenSAFELY tools. These tools in turn enable researchers with OpenSAFELY Data Access Agreements to write and execute code for data management and data analysis without direct access to the underlying raw pseudonymised patient data, and to review the outputs of this code. All code for the full data management pipeline from raw data to completed results for this analysis and for the OpenSAFELY platform as a whole is available for review at github.com/OpenSAFELY.

https://github.com/opensafely/covid-19-vaccine-breakthrough

## Abbreviations

(BMI): Body Mass index
(JCVI): Joint Committee Vaccination and Immunisation
(IMD): Index of Multiple Deprivation
(UK): United Kingdom

## Declarations

### Information governance and ethical approval

NHS England is the data controller; TPP is the data processor; and the key researchers on OpenSAFELY are acting with the approval of NHS England. This implementation of OpenSAFELY is hosted within the TPP environment which is accredited to the ISO 27001 information security standard and is NHS IG Toolkit compliant^[17,18]^; Patient data has been pseudonymised for analysis and linkage using industry standard cryptographic hashing techniques; all pseudonymised datasets transmitted for linkage onto OpenSAFELY are encrypted; access to the platform is via a virtual private network (VPN) connection, restricted to a small group of researchers; the researchers hold contracts with NHS England and only access the platform to initiate database queries and statistical models; all database activity is logged; only aggregate statistical outputs leave the platform environment following best practice for anonymisation of results such as statistical disclosure control for low cell counts^16^. The OpenSAFELY research platform adheres to the obligations of the UK General Data Protection Regulation (GDPR) and the Data Protection Act 2018. In March 2020, the Secretary of State for Health and Social Care used powers under the UK Health Service (Control of Patient Information) Regulations 2002 (COPI) to require organisations to process confidential patient information for the purposes of protecting public health, providing healthcare services to the public and monitoring and managing the COVID-19 outbreak and incidents of exposure; this sets aside the requirement for patient consent^17^. Taken together, these provide the legal bases to link patient datasets on the OpenSAFELY platform. GP practices, from which the primary care data are obtained, are required to share relevant health information to support the public health response to the pandemic, and have been informed of the OpenSAFELY analytics platform.

This study was approved by the Health Research Authority (REC reference 20/LO/0651) and by the LSHTM Ethics Board (reference 21863).

### Availability of data and materials

Access to the underlying identifiable and potentially re-identifiable pseudonymised electronic health record data is tightly governed by various legislative and regulatory frameworks, and restricted by best practice. The data in OpenSAFELY is drawn from General Practice data across England where TPP is the Data Processor. TPP developers (CB, RC, JP, FH, and SH) initiate an automated process to create pseudonymised records in the core OpenSAFELY database, which are copies of key structured data tables in the identifiable records. These are linked onto key external data resources that have also been pseudonymised via SHA-512 one-way hashing of NHS numbers using a shared salt. DataLab developers and PIs (BG, LS, CEM, SB, AJW, WH, DE, PI, and CTR) holding contracts with NHS England have access to the OpenSAFELY pseudonymised data tables as needed to develop the OpenSAFELY tools. These tools in turn enable researchers with OpenSAFELY Data Access Agreements to write and execute code for data management and data analysis without direct access to the underlying raw pseudonymised patient data, and to review the outputs of this code. All code for the full data management pipeline—from raw data to completed results for this analysis—and for the OpenSAFELY platform as a whole is available for review at github.com/OpenSAFELY.

## Authors’ contributions

Contributions are as follows:

Conceptualisation: RM, CTR, KB, RME, LS, BG, BM, HJC, SJWE, KK, DH, KR

Funding acquisition: LS, BG, RME

Methodology: AG, HC, WH, EW, HMD, AW, LF, CA, LH, CM, BMK, RC, JM, AM, KB,AS,CR,AR,LF,LT, RM, AW, HF, SE, LS, BG

Formal analysis: AG, HC

Codelists: RM, LT, AS, AJW, CM, BG, WJH, SB, AM

Software: AW, CB, JC, DE, PI, CM, WH, BN, SB, HC, ND, RC, JP, FH, SH

Visualisation: AG, HC, WH, EW

Writing - original draft: AG

Writing-review & editing: ALL

Information governance: CB LS BG AM

## Acknowledgements

We are very grateful for all the support received from the TPP Technical Operations team throughout this work, and for generous assistance from the information governance and database teams at NHS England / NHSX.

## Guarantor

BG is guarantor.

## References

1. Coronavirus (COVID-19) in the UK. In: GOV.UK. https://coronavirus.data.gov.uk/details/vaccinations. Accessed 9 Sep 2021

2. Bernal JL, Andrews N, Gower C, et al (2021) Effectiveness of Covid-19 Vaccines against the B.1.617.2 (Delta) Variant. New England Journal of Medicine 385:585–594

3. Lopez Bernal J, Andrews N, Gower C, et al (2021) Effectiveness of the Pfizer-BioNTech and Oxford-AstraZeneca vaccines on covid-19 related symptoms, hospital admissions, and mortality in older adults in England: test negative case-control study. BMJ 373:n1088

4. OpenSAFELY. https://www.opensafely.org/. Accessed 20 Oct 2021

5. Emergency use ICD codes for COVID-19 disease outbreak. https://www.who.int/classifications/classification-of-diseases/emergency-use-icd-codes-for-covid-19-disease-outbreak. Accessed 20 Oct 2021

6. UK Health Security Agency (2020) COVID-19: the green book, chapter 14a. https://www.gov.uk/government/publications/covid-19-the-green-book-chapter-14a. Accessed 20 Oct 2021

7. COVID-19 Vaccination Uptake Reporting Specification. PRMIS. https://www.nottingham.ac.uk/primis/covid-19/covid-19.aspx. Accessed 20 Jun 2021

8. Cook C, Patel NJ, D’Silva KM, et al (2021) Clinical characteristics and outcomes of COVID-19 breakthrough infections among vaccinated patients with systemic autoimmune rheumatic diseases. Ann Rheum Dis. https://doi.org/10.1136/annrheumdis-2021-221326

9. Keehner J, Horton LE, Binkin NJ, Laurent LC, Pride D, Longhurst CA, Abeles SR, Torriani FJ (2021) Resurgence of SARS-CoV-2 Infection in a Highly Vaccinated Health System Workforce. N Engl J Med 385:1330–1332

10. Oster Y, Benenson S, Yochi Harpaz L, Buda I, Nir-Paz R, Strahilevitz J, Cohen MJ (2021) Association Between Exposure Characteristics and the Risk for COVID-19 Infection Among Health Care Workers With and Without BNT162b2 Vaccination. JAMA Netw Open 4:e2125394

11. Antonelli M, Penfold RS, Merino J, et al (2021) Risk factors and disease profile of post-vaccination SARS-CoV-2 infection in UK users of the COVID Symptom Study app: a prospective, community-based, nested, case-control study. Lancet Infect Dis. https://doi.org/10.1016/S1473-3099(21)00460-6

12. Hippisley-Cox J, Coupland CA, Mehta N, et al (2021) Risk prediction of covid-19 related death and hospital admission in adults after covid-19 vaccination: national prospective cohort study. BMJ 374:n2244

13. Day M (2021) Covid-19: Stronger warnings are needed to curb socialising after vaccination, say doctors and behavioural scientists. BMJ. https://doi.org/10.1136/bmj.n783

14. Polack FP, Thomas SJ, Kitchin N, et al (2020) Safety and Efficacy of the BNT162b2 mRNA Covid-19 Vaccine. N Engl J Med 383:2603–2615

