## Supplementary material for "Describing the population experiencing COVID-19 vaccine breakthrough following second vaccination in England: A cohort study from OpenSAFELY"

### Chronic kidney disease

To demonstrate the public health burden (rather than likely causal mechanisms), within patients with chronic kidney disease (CKD), Table S1 shows both crude and adjusted rates of positive SARS-CoV-2 swab test, COVID-19 related hospital admissions, COVID-19 related critical care admissions and COVID-19 related death, broken down by CKD stage. Incidence rates for each outcome were estimated by dividing the count by person-years, with 95% confidence intervals. Crude and adjusted rates were also calculated using cox regression and show that even after adjustment, rates remain very high. These results highlight that fully vaccinated individuals with kidney disease are at far higher risk of serious outcomes when infected with COVID-19 than individuals without kidney disease.

**Table S1:** Number of fully vaccinated (2 doses + 2 weeks) patients with chronic kidney disease by stage, in OpenSAFELY-TPP, and associated crude and adjusted rates of positive SARS-CoV-2 swab test, COVID-19 related hospital admissions, COVID-19 related critical care admissions and COVID-19 related death, broken down by CKD stage.

| **Population/Outcome** | | | **Kidney disease** | | | | |
|---|---|---|---|---|---|---|---|
| | | | **No CKD** | **Stage 3a** | **Stage 3b** | **Stage 4** | **Stage 5** |
| **Fully vaccinated** | **Count*** | | 14622275 | 527255 | 205755 | 51035 | 5505 |
| | **Follow-up time, medium days (IQR)** | | 147 (103-177) | 187 (173-201) | 192 (178-205) | 191 (174-204) | 184 (164-200) |
| | **Tests conducted (%)** | **0** | 59 | 61 | 60 | 55 | 37 |
| | | **1** | 17 | 14 | 13 | 12 | 12 |
| | | **2** | 7 | 6 | 6 | 7 | 8 |
| | | **3+** | 17 | 18 | 21 | 26 | 44 |
| **Positive SARS-CoV-2 test** | **Positivity rate, %** | | 2.00 | 1.80 | 1.77 | 1.77 | 1.42 |
| | **Events* (PYs)** | | 557715 (5474016) | 12135 (267915) | 4440 (106744) | 1275 (25816) | 215 (2607) |
| | **Incidence rate per 10,000 person-years (95% CI)** | | 101.88 (101.75-102.02) | 45.29 (44.88-45.7) | 41.59 (40.97-42.21) | 49.35 (47.99-50.75) | 82.86 (77.41-88.69) |
| | **Hazard ratio** | | 1.00 | 0.41 (0.40-0.41) | 0.37 (0.36-0.38) | 0.44 (0.42-0.47) | 0.76 (0.66-0.86) |
| | **Age-adjusted hazard ratio** | | 1.00 | 0.79 (0.77-0.80) | 0.80 (0.78-0.83) | 0.95 (0.90-1.00) | 1.29 (1.13-1.48) |
| **COVID-19 related hospital admission** | **Events* (PYs)** | | 11865 (5513255) | 2040 (267918) | 1270 (106641) | 465 (25788) | 80 (2615) |
| | **Incidence rate per 10,000 person-years (95% CI)** | | 2.15 (2.13-2.17) | 7.61 (7.44-7.78) | 11.89 (11.56-12.23) | 18.11 (17.29-18.97) | 30.97 (27.72-34.61) |

| | | | | | | |
|---|---|---|---|---|---|---|
| | **Hazard ratio** | 1.00 | 3.00 (2.86-3.14) | 4.61 (4.35-4.88) | 7.08 (6.45-7.77) | 12.47 (10.02-15.52) |
| | **Age-adjusted hazard ratio** | 1.00 | 1.84 (1.75-1.93) | 2.56 (2.41-2.72) | 3.88 (3.53-4.27) | 8.01 (6.43-9.97) |
| **COVID-19 related critical care admission** | **Events* (PYs)** | 815 (5514965) | 105 (268206) | 55 (106814) | 25 (25846) | 10 (2625) |
| | **Incidence rate per 10,000 person-years (95% CI)** | 0.15 (0.14-0.15) | 0.4 (0.36-0.44) | 0.51 (0.45-0.59) | 1.04 (0.86-1.27) | 3.43 (2.46-4.78) |
| | **Hazard ratio** | 1.00 | 2.45 (2.00-3.00) | 3.21 (2.44-4.23) | 6.57 (4.47-9.64) | 21.81 (11.30-42.07) |
| | **Age-adjusted hazard ratio** | 1.00 | 1.92 (1.55-2.38) | 2.40 (1.55-2.38) | 4.88 (3.29-7.23) | 17.52 (9.05-33.90) |
| **COVID-19 related death** | **Events* (PYs)** | 2370 (5554595) | 690 (269616) | 500 (107352) | 215 (25976) | 30 (2639) |
| | **Incidence rate per 10,000 person-years (95% CI)** | 0.43 (0.42-0.44) | 2.55 (2.46-2.65) | 4.67 (4.46-4.88) | 8.28 (7.73-8.86) | 11.37 (9.47-13.65) |
| | **Hazard ratio** | 1.00 | 4.23 (3.88-4.61) | 7.36 (6.68-8.12) | 13.27 (11.53-15.28) | 19.53 (13.62-28.00) |
| | **Age-adjusted hazard ratio** | 1.00 | 1.68 (1.54-1.84) | 2.33 (2.10-2.58) | 3.98 (3.45-4.60) | 8.08 (5.63-11.60) |
